# Non-invasive Real-time Detection of Potassium level Changes in Skeletal Muscles during Exercise by Magnetic Resonance Spectroscopy

**DOI:** 10.1101/2024.09.30.24314637

**Authors:** Etienne Roesli, Hedvika Haindrich Primasová, Marc Thiede, Laila-Yasmin Mani, Lena V. Gast, Armin M. Nagel, Bruno Vogt, Peter Vermathen

## Abstract

**Introduction:** Potassium is essential in cellular functions, with specific importance in muscle activity and cardiovascular health. It is the main intracellular cation in the human body with 70% located in muscle. Traditional methods to measure potassium levels are invasive and lack specificity for intracellular concentrations. Recently, non-invasive in vivo investigation of K+ ion homeostasis has become feasible by using ^39^K Magnetic Resonance Imaging (MRI) and MR spectroscopy (MRS) at ultrahigh magnetic fields. However, studies demonstrating the sensitivity of ^39^K MRI or MRS to detect potassium alterations in disease or upon intervention are sparse. This study utilizes ^39^K MRS to non-invasively track real-time intramuscular potassium changes during exercise, providing an assessment of potassium dynamics and explores the potential for technical artifacts in the measurements.

**Methods:** Five healthy subjects (three males, two females) were recruited to perform standardized dynamic knee extensions inside a 7T MR scanner. Potassium levels were measured using a ^39^K MRS protocol that included periods of rest, moderate, and heavy exercise followed by recovery. Additionally, possible measurement artifacts due to muscle movement or changes in coil position relative to the thigh were evaluated using K MRS and H MRI monitoring in separate sessions.

**Results:** The study revealed a consistent decrease in potassium levels during both moderate and heavy exercise, with an average decrease of 5-6%. These changes were rapidly detectable and were reversed upon cessation of exercise, indicating effective in vivo monitoring capability. Possible experimental artifacts were investigated, and the results suggested not to be responsible for the detected potassium changes during exercise. The results of the non-localized ^39^K MRS measurements during exercise correlated well with expected physiological changes based on previous literature.

**Discussion:** The application of ^39^K MRS provides a valuable non-invasive tool for studying potassium dynamics in human skeletal muscle. This technique could enhance our understanding of muscle physiology and metabolic disorders. The ability to measure these changes in real time and non-invasively highlights the potential for clinical applications, including monitoring of diseases affecting muscle and cellular metabolism.

## Introduction

Potassium and sodium play vital roles in numerous cellular processes. Potassium has been classified as “a nutrient of public health concern” by the U.S. FDA because of under-consumption. Low potassium intake associates with cardiovascular disease and mortality, while beneficial effects of higher potassium intake have been demonstrated [1–3]. Since serum or urine potassium levels correlate poorly with tissue potassium because the majority of potassium is inside the cells, its specific determination in different organs would be very valuable. Existing methods are invasive, associated with radiation exposure and lack spatial resolution [4–6]. However, while Na-MR imaging has been proven to be clinically applicable, non-invasive in vivo determination of ^39^K was not possible until very recently. By using ^39^K MR spectroscopy (MRS) and MRI at ultrahigh magnetic fields, it has recently been convincingly demonstrated for the first time that a non-invasive method for in vivo investigation of the important K ion homeostasis and of normal cell membrane function i.e., Na - K - ATPase function in humans, has become feasible [7–13].

It is well established that the distribution of potassium within the body is of great importance, not only in terms of the gradient between extracellular and intracellular concentrations, but also in relation to the different concentrations present in different tissues. The gradient of potassium concentration across the cellular membrane is essential for its function, especially for the membrane potential. The average intracellular concentration is approximately 125 mM, while the extracellular concentration ranges from 3.5 to 5.4 mM [14]. Furthermore, potassium is not distributed homogeneously within body tissues. Skeletal muscle tissue contains the majority of the body’s potassium, with approximately 68% [14]. Given that skeletal muscle tissue makes up about 40% of the body’s weight, it plays a crucial role in maintaining plasma potassium concentration at an equilibrium state. Following ingestion, up to 80% of the ingested K+ is transported to the intracellular space. Skeletal muscle tissue, liver and kidneys are the primary organs responsible for this process [14].

Exercise has a profound effect on the homeostasis of potassium. During exercise, muscle fibres depolarise at a high frequency, resulting in a loss of potassium from muscle cells. In this situation, the transport proteins for potassium are unable to maintain an equilibrium state, therefore the intracellular concentration lowers, whereas the plasma and interstitial concentrations increase. Changes of potassium concentration during exercise have been investigated in detail with invasive methods in the past. These studies showed that total muscle potassium concentration decreases by approximately 6-10% during exhaustive exercise [15,16]. In the intracellular space, the potassium concentration decreases even more, by approximately 20%. However, some of the potassium is temporarily stored in the interstitial space, where the potassium concentration increases to 9 mM and above [17].

Conversely, following the cessation of exercise, potassium is shifted back to the muscle tissue due to the highly activated Na+ K+ pump. This process has been described to be composed of two phases, an initial fast component and a second slower one. The first phase is designed to correct the high potassium in the plasma back to physiological levels. It starts immediately after the cessation of the exercise and holds on for seconds to a few minutes. The second phase is most often characterized by an undershoot of plasma potassium levels. Due to this, the reuptake of potassium back to the muscle is limited as it would be life threatening, if the plasma potassium decreases too much. This second phase however is not well understood [18].

In the past, only a few applications of ^39^K MRI were described in the literature [7–13]. The main reason for this is the very low NMR signal intensity of ^39^K, which is due to a 2000 times lower NMR sensitivity and a 1000 times lower concentration in human tissue than protons [19]. Nevertheless, with the increasing availability of MR scanners with ultra-high field strengths of 7 Tesla and beyond, in vivo investigations of X-nuclei with low sensitivities have become feasible.

While the ability to measure potassium concentrations in vivo using MRI, MRS, or magnetic resonance spectroscopic imaging (MRSI) has been shown convincingly in several studies, to our knowledge only a single study has shown that changes in potassium concentration can also be measured [13]. In a first proof of concept study, we therefore aim at determining the feasibility of detecting potassium changes in skeletal muscles using ^39^K-MRS during exercise. A recent ^39^K-MRI study in human calf muscle tissue determined a coefficient of variation between scan and re-scan of 6% for ^39^K-content estimation [11]. Based on this study we hypothesized that a potassium change of 6-10 % in quadriceps muscles of the thigh during exercise can be monitored by ^39^K-MRS.

The study by L. Gast et al. [13] reported a non-significant tendency for a decrease in the apparent calf muscle potassium concentration directly after exercise, but only in subjects with a high increase in serum creatine kinase. As the authors noted in their discussion, the observed decrease in potassium concentration could have been greater had the exercise and potassium MRI been performed within a shorter time gap. Consequently, the objective of the present study was to conduct measurements on quadriceps muscles of the thigh during exercise and recovery using non-localized MR spectroscopy, with a high temporal resolution of approximately one minute, and to determine if adaptations of the potassium concentration in the muscle can be followed in real-time.

In the main experiment of the study, five subjects engaged in a series of exercise routines performed within the MR-scanner, with the progression of potassium levels being monitored simultaneously.

Several technical aspects could potentially lead to the generation of artefacts during the MRS measurements during exercise. First, although the coil was fixed as securely as possible, the exercise could cause coil slipping in relation to the thigh. This may lead to an altered muscle mass within the sensitive volume of the coil and therefore to an artificially changed potassium signal. Second, the contraction of the muscle may also alter the total muscle mass within the sensitive volume of the coil, resulting in a similar consequence. To address this issue, three additional subjects were recruited to investigate and potentially exclude the possibility of artificial findings.

## Methods

### Subjects

A total of five healthy and young students (three males and two females, S1-S5) consented to participate in the initial phase of the study (the main experiment). The inclusion criterion was that they were able to perform knee extensions for a duration of 20 minutes. The level of sport activity was evaluated using the Tegner Activity Scale (TAS) [20]. According to the survey, the participants had an average score of 5 on the TAS. The mean age of the participants was 23.4±1.5 years, with a mean body mass index of 23.0±2.4 kg/m^2^. Subject S5 consented to further measurements following the main experiment, while three additional subjects (all male, S6-S8) were recruited solely for the purpose of investigating potential artefacts, thus resulting in a total of four subjects for this specific issue. Exclusion criteria included the presence of tattoos in the sensitive area of the coil, metal components within the body and other general contraindications for MRI. The measurements were approved by the local ethics committee and all subjects provided written informed consent. It should be noted that the study population is biased, as all participants were young adults, with the majority being male.

One participant (S4) regularly takes Lisinopril, an ACE-inhibitor, which influences the potassium homeostasis in the body [21]. Despite a thorough search of the literature, no evidence was found indicating whether this type of medication influences potassium concentration in skeletal muscle during exercise. However, ACE-inhibitors may have an effect on plasma K+ concentration during exercise [22].

### MR Setup

Exercise and MR examinations were performed in a 7T MR scanner (Terra, Siemens, Erlangen, Germany). A H/ K double tuned transmit/receive surface coil (RAPID Biomedical, Rimpar, Germany) with an 18 cm diameter for ^39^K was utilised to record the signal. The coil design included two reference tube holders that were filled with bottles containing chemically shifted potassium solutions (chemical shift reagent Tm-DOTP) for quantification and for monitoring the coil positioning by MRI. The coil was positioned directly on the right thigh of the subject and both coil and thigh were fixed as securely as possible to minimize unintended movement during exercise. Fig. 1 shows the setup in the MR scanner. A leg support was used, and the knees were bent approximately 28° degrees at rest. Dynamic knee extensions to stretched position (0°) activating the quadriceps were performed against a stretch band that was fixed at the right ankle. For this initial feasibility study, the exercise intensity was not quantified.

**Figure 1:**
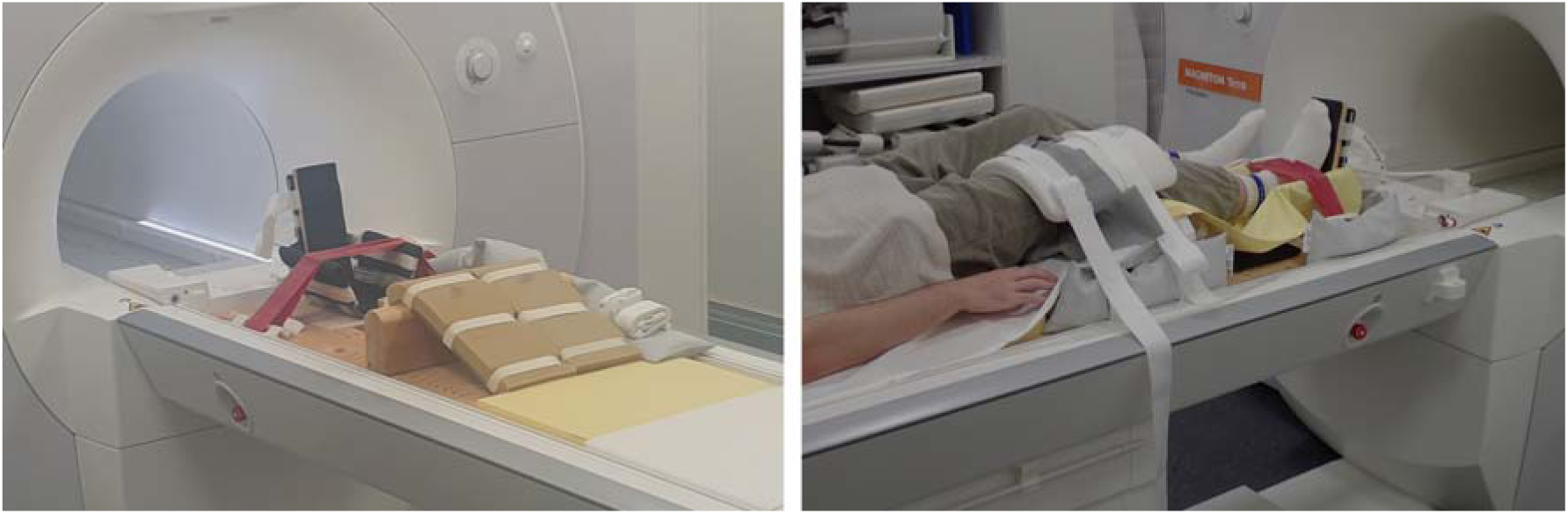
Experimental setup for the exercise investigations in the 7T MR scanner. Left: Self-made simple MR compatible exercise unit with upper and lower leg support and stretch band. Right: Positioning of a volunteer on the exercise unit and positioning and fixing of the surface coil on the thigh.

### Exercise intervention

#### Main experiment

The exercise protocol is visualized in Fig. 2. The baseline potassium signal was measured for a period of nine minutes (10 spectra of 54 s each). Following the baseline measurement, the subjects were instructed to perform moderate periodic knee extensions with stretching and relaxation against the stretch band for 13.5 min (15 x 54 s) and then to perform heavy knee extensions for another 4.5 min (5 x 54 s). The levels “moderate” and “heavy” were determined subjectively by the participants and may have varied during the exercise period due to the challenging task of splitting up the force for such a long period of time. After the phase of heavy knee extensions, all subjects reported a state of exhaustion or near-exhaustion. Subsequently, the recovery phase was again monitored for nine minutes (10 x 54 s).

**Figure 2:**
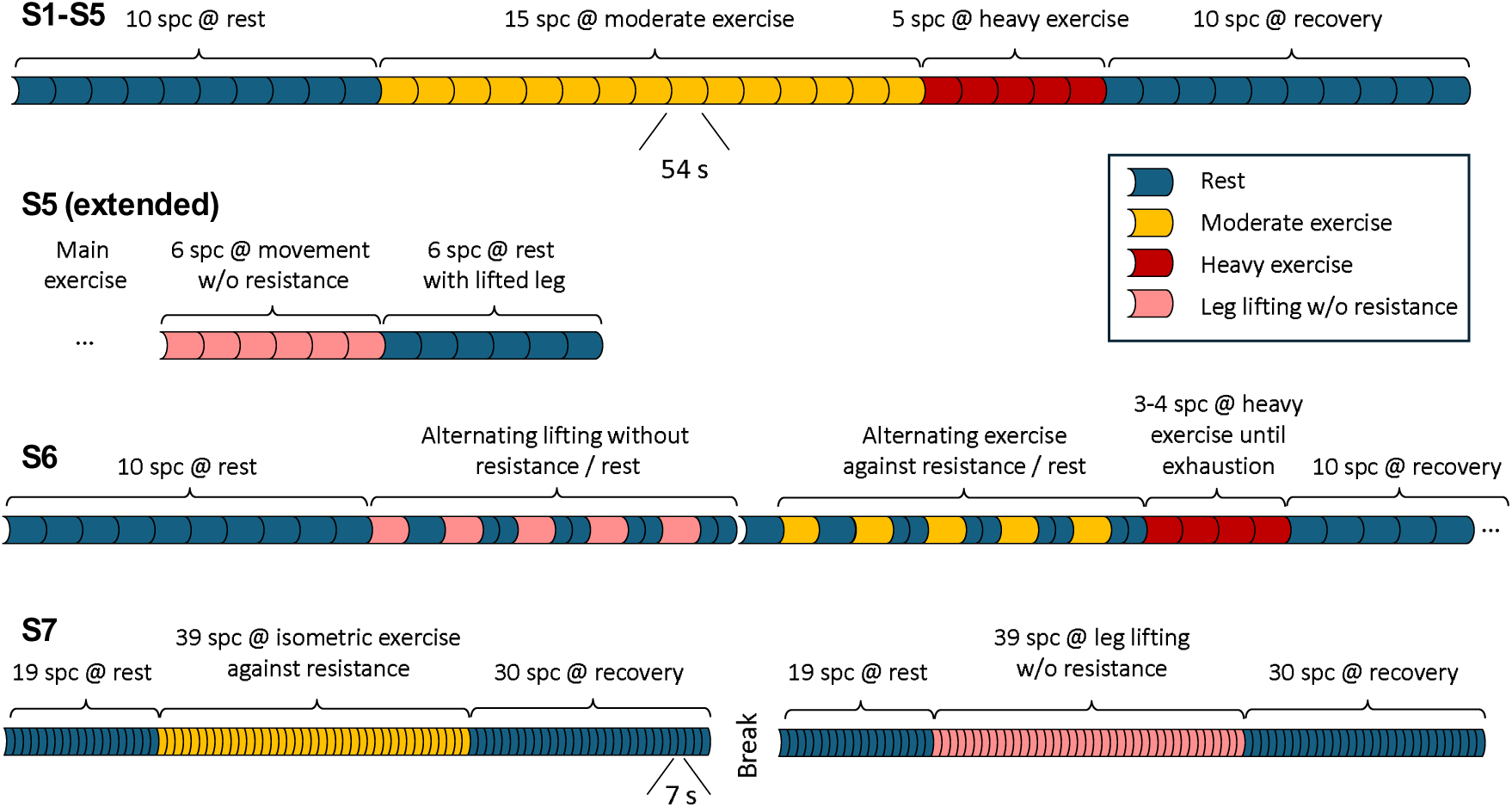
Top: Experimental protocol for the main exercise study (S1-S5). Below: Individual protocols for the additionally performed validation MR Spectroscopy measurements on three subjects, S5, S6 and S7.

#### Investigation of possible artefacts

Four subjects were assigned to perform exercises that differed from the protocol of the main experiment. Each subject had a separate protocol to investigate different technical aspects of the measurement.

To determine the impact of thigh muscle movement by lower leg extensions without resistance on the detected potassium signal, subject S5 performed knee extensions for 5.4 min (6 spectra of 54 s each) without stretch band following the main experiment (Fig. 2). Because the power duty of the muscle is much lower, the potassium depletion was expected to be less pronounced as with stretch band. With the same rationale to determine the impact of altered lower leg position on the thigh position in relation to the coil the subject’s lower leg was subsequently raised on a support so that the knee was stretched at rest. The ^39^K-signal was once again measured for 5.4 min.

Subject S6 performed – after an initial resting period – alternating exercise and recovery for 54 s each five times in a row without resistance of the stretch band. Afterwards, the measurement was repeated with the stretch band (with resistance) again alternating exercise for 54 s, followed by 54 s recovery (split to 2 x 27 s for the last four repetitions), five times repeated (Fig. 2). The rationale of this experiment was to further investigate the kinetics of ^39^K depletion and recovery. Finally, the subject was encouraged to exercise until exhaustion, which lasted 3-4 min. The recovery phase was monitored as noted above.

Subject S7 performed isometric leg lifting first without the stretch band and afterwards isometric exercise against resistance of the stretch band. This time, all spectra were measured with a time resolution of 6.75 s using only 32 acquisitions. The objective of this investigation was first to eliminate the periodic lower leg movement, and second, to potentially detect a transition period of potassium changes after exercise onset or cessation. The baseline was measured for a duration of 135 s (20 x 6.75 s). Subsequently, the subject lifted the leg without the stretch band for a period of 270 s (40 x 6.75 s). Then the recovery period was measured for a duration of 210 s (30 x 6.75 s). The same protocol was then replicated against resistance with the addition of the stretch band.

To directly detect potential thigh movement and contraction during lower leg exercise, fast H-MRI monitoring was performed in subject S8 as described below (in “MR imaging during Exercise”) while performing the same knee extensions as for the real-time ^39^K investigations. S8 performed periodic knee extensions with stretching and relaxation against the stretch band for 12 s (performing 6 periodic extensions while recording 50 MR-images) followed by a 12 s rest period (again monitored by 50 MR-images). The experiment was repeated without the stretch band (performing 8 periodic extensions).

### MR Acquisitions

#### MR spectroscopy

The coil positioning close to vastus lateralis and rectus femoris was verified on localizer MR images acquired in sagittal, coronal, and transverse orientations. For ^39^K-MR spectroscopy, an unlocalized simple free induction decay (FID) sequence was employed for maximum signal yield to allow for fast acquisition in real-time during exercise, i.e., the sensitivity profile of the surface coil determined the localization of the detected signal. Parameters of the FID sequence included a pulse length of 0.5 ms, a 0.35 ms interval between the pulse and acquisition, a repetition time (TR) of 210 ms, sweep width of 2500 Hz, 256 data points, and 256 acquisitions, resulting in a total duration of 54 s for the main experiment. Prior to the measurement ^39^K frequency was adjusted; no shimming was performed. Measurements were taken continuously in the main study while the subject was engaged in baseline, moderate and heavy exercise as well as during recovery. For the additionally performed MRS measurements, the same sequence parameters were used, only the number of acquisitions was varied.

#### Spectral processing

The acquired spectra were processed using Java-based magnetic resonance user interface (jMRUI) [23]. Processing included zero-filling to 1024 points, 5 Hz Lorentzian line broadening, and phase correction. The muscle spectra demonstrated broad signal contributions from quadrupolar splitting of the potassium signal [8]. To improve robustness of the spectral fitting contributions these broad components which have very short T2*-values were minimized by eliminating the first 16 data points of the acquisition. Spectral fitting was performed using jMRUI AMARES (advanced method for accurate, robust, and efficient spectral fitting) [23]. Lorentzian lines were used for fitting the tissue ^39^K and the chemically shifted ^39^K-reference lines. The frequency shift of the reference line was limited not to overlap with the tissue ^39^K resonance. Both fitted peak amplitude (which can be determined very precisely) and peak area (which is less vulnerable to potential line broadening effects) of the potassium resonances were considered for further analysis. The values from all spectra obtained for each subject were normalized to the mean value of the baseline period to allow for comparisons between subjects. The values were also related to the value of the chemically shifted reference line, which provides a constant signal even during movement. The relative potassium changes were compared within each subject longitudinally between the different phases (baseline, moderate exercise, heavy exercise and recovery) and between subjects.

#### MR imaging during Exercise

To investigate any movement or shape changes of the thigh during exercise, potentially changing total muscle area within the sensitive volume during exercise, MR images were acquired with high temporal resolution of 250 ms. A 2D TrueFISP (tfi2d) sequence was applied in transverse orientation selecting a single cross section in the middle of the fixed quadriceps muscle (FOV 230×230 mm , slice thickness 5 mm, repetition time (TR) 3.23 ms, echo time (TE) 1.62 ms, flip angle 46°, resolution 129×90, acquisition time 250 ms).

The MR images were analyzed using the Medical Image Processing, Analysis, and Visualization (MIPAV) application [24]. Muscle contour plots and the estimated tissue within the sensitive region of the coil (obtained from preceding phantom measurements) were used to determine the enclosed muscle tissue area.

#### Statistics

Assuming that the data follow a statistical normal distribution, t-tests were applied. Unpaired t-tests were used to investigate the significance of changes between different exercise-phases within each subject using the repeatedly determined ^39^K values of the different phases. Paired t-tests were applied to investigate the significance of mean potassium changes for each exercise-phase across subjects. A limitation of this protocol is the low number of longitudinal data points especially during the period of heavy exercise as well as the low number of subjects following the same exercise protocol.

## Results

### Main experiment

A signal-to-noise ratio (SNR) of >50 was obtained for the individual unlocalized ^39^K-spectra with a duration of 54 s. Fig. 3 show representative spectra for one subject. Furthermore, inspection of the spectra in Fig. 3 also demonstrated different peak intensities for the different exercise phases.

**Figure 3:**
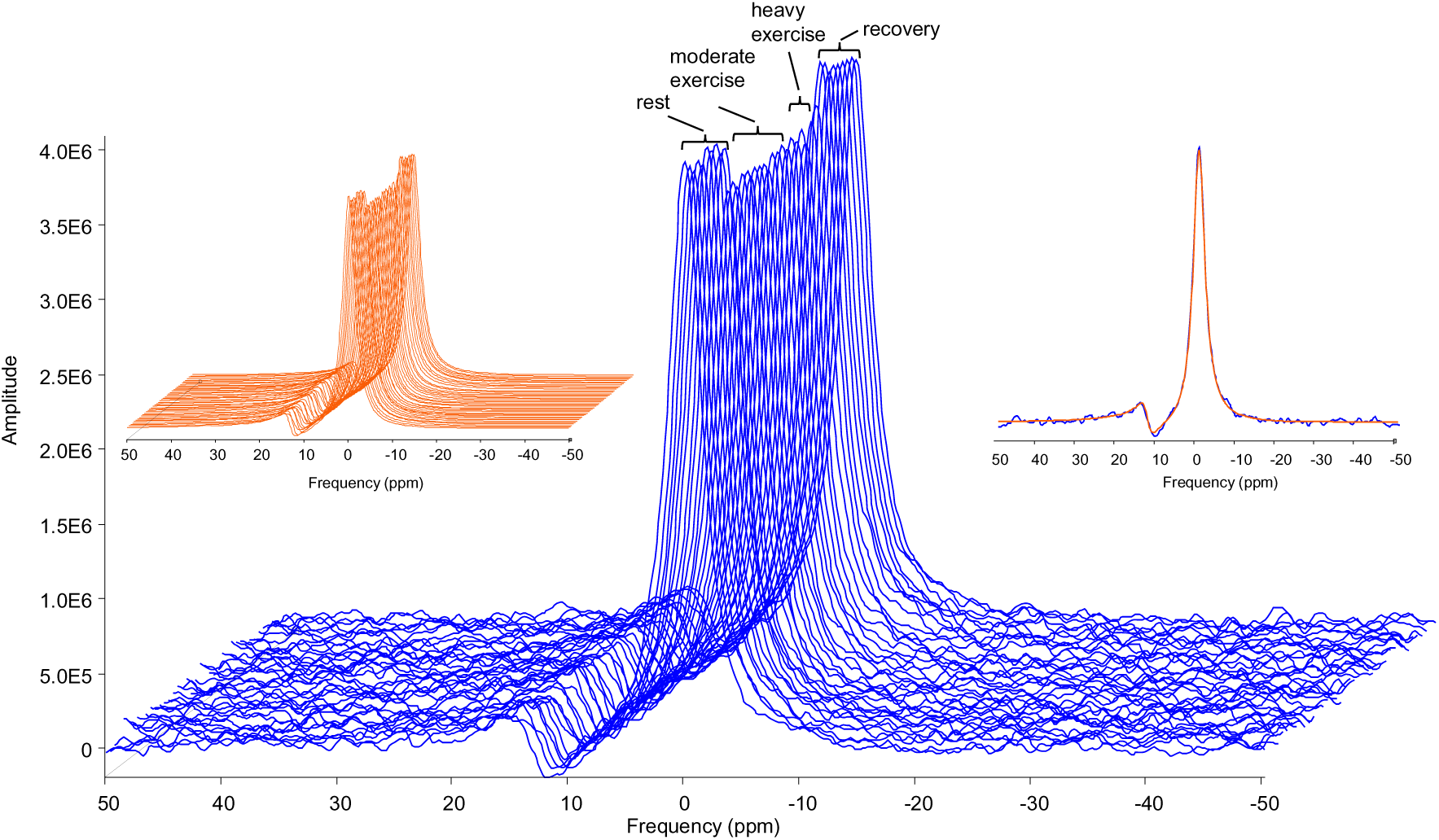
Main exercise study. Example K spectra from subject S5. Different exercise periods are indicated. The signal amplitudes are different depending on exercise periods. Left insert: Corresponding fitted spectra using jMRUI [23]. Right insert: Single spectrum with fitted spectrum overlaid. The peak at ∼12 ppm originates from the chemically shifted K signal of the reference phantom.

Figure 4a) illustrates the quantitative analysis of the individual spectra (relative to the mean value of the baseline period) for the five subjects investigated applying the same protocol. The mean values and standard deviations for each period of the protocol for all five subjects who participated in the main experiment are presented in Figs. 4b and 4c for fitted peak-height and peak-area, respectively. The corresponding numerical values and statistical results for the comparisons of the different phases within and across subjects are presented in Table 1. Analysis of peak area and height yielded similar results, validating that potential line-broadening during exercise was not responsible for the detected K-level changes.

**Figure 4:**
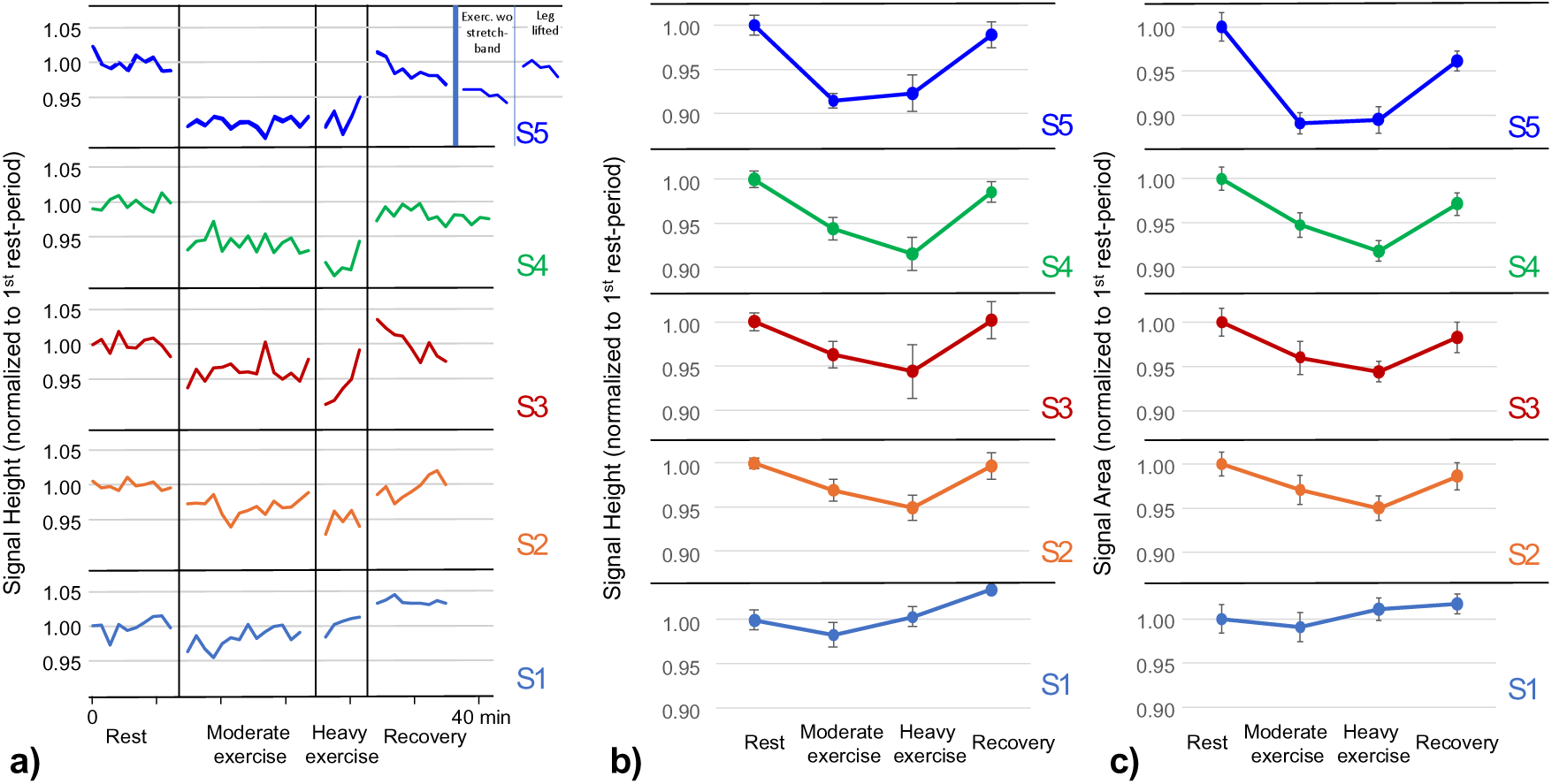
a) Quantitative analysis of the fitted peak-height of the individual spectra (relative to the mean value of the baseline period) for the five subjects who participated in the main experiment applying the exercise protocol provided in Fig 2 (top). The plot for S5 illustrates in addition the results of the extended measurement without stretch band and with lifted leg. b, c) Mean values and standard deviations for each period of the protocol for the five subjects for fitted peak-height and peak-area, respectively.

**Table 1:** Average values and standard deviations for each period of the protocol for the five subjects for fitted peak-height and peak-area, respectively. Right column: Mean value over all subjects for each exercise phase. Significant differences between the phases are indicated.

| Height (normalized to first rest period) |  |  |  |  |  |  |
| --- | --- | --- | --- | --- | --- | --- |
|  | S1 | S2 | S3 | S4 | S5 | Mean |
| Rest | 1.00±0.01 | 1.00±0.01 | 1.00±0.01 | 1.00±0.01 | 1.00±0.01 | 1.00±0.00 |
| Moderate Exercise | 0.98±0.01<sup>*§</sup> | 0.97±0.01<sup>*§</sup> | 0.96±0.02<sup>*§</sup> | 0.94±0.01<sup>*§</sup> | 0.91±0.01<sup>*§</sup> | 0.95±0.03<sup>**§§</sup> |
| Heavy Exercise | 1.00±0.01<sup>§</sup> | 0.95±0.01<sup>*§</sup> | 0.94±0.03<sup>*§</sup> | 0.92±0.02<sup>*§</sup> | 0.92±0.02<sup>*§</sup> | 0.95±0.03<sup>**§§</sup> |
| Recovery | 1.03±0.00<sup>*</sup> | 1.00±0.02 | 1.00±0.02 | 0.98±0.01<sup>*</sup> | 0.99±0.01 | 1.00±0.02 |

| Area (normalized to first rest period) |  |  |  |  |  |  |
| --- | --- | --- | --- | --- | --- | --- |
|  | S1 | S2 | S3 | S4 | S5 | Mean |
| Rest | 1.00±0.02 | 1.00±0.01 | 1.00±0.02 | 1.00±0.01 | 1.00±0.02 | 1.00±0.00 |
| Moderate Exercise | 0.99±0.02<sup>§</sup> | 0.97±0.02<sup>*§</sup> | 0.96±0.02<sup>*§</sup> | 0.95±0.01<sup>*§</sup> | 0.89±0.01<sup>*§</sup> | 0.95±0.04<sup>**§§</sup> |
| Heavy Exercise | 1.01±0.01 | 0.95±0.01<sup>*§</sup> | 0.94±0.01<sup>*§</sup> | 0.92±0.01<sup>*§</sup> | 0.89±0.02<sup>*§</sup> | 0.94±0.04<sup>**§§</sup> |
| Recovery | 1.02±0.01<sup>*</sup> | 0.99±0.02 | 0.98±0.02<sup>*</sup> | 0.97±0.01<sup>*</sup> | 0.96±0.01<sup>*</sup> | 0.98±0.02 |

\*: p-value < 0.05 (unpaired t-test) comparing individual <sup>39</sup>K-levels of moderate, heavy exercise, and recovery with levels at rest
\*\* : p-value < 0.05 (paired t-test) comparing mean <sup>39</sup>K-levels of all 5 subjects of moderate, heavy exercise, and recovery with levels at rest
§: p-value < 0.05 (unpaired t-test) comparing individual <sup>39</sup>K-levels of moderate and heavy exercise with recovery levels
§§: p-value < 0.05 (paired t-test) comparing mean <sup>39</sup>K-levels of all 5 subjects of moderate, heavy exercise, and recovery with levels at rest

### Baseline vs Exercise

The analysis of the peak height of the central ^39^K-resonance revealed relatively stable values for the series of spectra during rest and recovery periods with standard deviations of less than 1.5% (Fig. 4a). The potassium signal decreased significantly in all five subjects during moderate exercise and in 4 of 5 subjects during heavy exercise compared to baseline. The decline was immediately apparent following the commencement of exercise (Fig. 4a). The minimum and maximum decreases were 2% and 9%, respectively, with a mean significant decrease of 4.5% during moderate and of 5.3% during heavy exercise. No consistent difference was detected between moderate and heavy exercise. The results of the analysis of the peak area were highly consistent (Fig. 4c and Table 1). The analysis of the muscle ^39^K resonance in relation to the ^39^K reference line revealed an even greater significant reduction during moderate and heavy exercise of 9.6% and 12.1%, respectively. However, due to low reference signal intensities the values for the series of spectra during rest and recovery periods showed a much larger standard deviation of 9.7% and 7.3%, respectively and the results are therefore subject to large errors.

In previous studies, the observed decrease in biopsies at maximum exercise was in the range of 6% to 10% [15, 16]. Our results are consistent with these findings.

### Exercise vs Recovery

As anticipated, potassium levels increased significantly back to the values at rest following the cessation of exercise due to a shift back to the muscle tissue [18]. This effect was observed in all subjects (Table 1). As for the ^39^K-signal decay after commencement of exercise, the potassium increase appeared immediately following the cessation of exercise (Fig. 4a). Compared to moderate and heavy exercise, the mean ^39^K-peak height increased significantly by 4.9% and 5.7%, respectively. In relation to the internal reference, the signal increased significantly compared to moderate and heavy exercise by 11.2% and 14.3%, respectively (again however, with much higher variance).

No significant differences were observed between values at rest and during recovery.

### Investigation of possible artefacts

As previously stated, four subjects underwent an altered exercise protocol in order to ascertain whether the observed decrease or increase in potassium signal was a consequence of potassium loss in the muscle or a result of the movement itself.

S5 performed leg extensions without the application of the stretch band, immediately following the recovery phase of the normal main experiment (Fig. 4a). In comparison to the preceding recovery period, the ^39^K-peak height was significantly reduced by 3.4% during exercise without stretch band, while the signal was 7.6% and 6.7% lower in this subject during moderate and heavy exercise, respectively against the stretch band. The following experiment raising the subject’s lower leg on a support so that the knee was stretched at rest resulted in similar ^39^K-values compared to values at rest or during recovery (Fig. 4a). The mean peak amplitude of this period did not differ from the baseline value of this subject.

S6 performed alternating exercise and recovery for 54 s each with and without stretch band as described in the methods section. The progression of the ^39^K signal height is illustrated in Fig. 5. During the periodic movement-rest periods without stretch band, the peak heights decreased only slightly during movement by less than 2% compared to the baseline period and increased immediately during the periodic rest periods. In contrast, during the periodic movement-rest periods with the stretch band, the signal decreased by 5.1% on average in relation to the baseline and increased immediately during the periodic rest periods, towards the end even above the baseline level. During the subsequent phase of heavy exercise, the potassium signal dropped by 3%, similar to the observations in the other subjects. At cessation of the heavy exercise part, which was – in contrast to S1-S5 determined by exhaustion - the signal increased almost instantly to 10% above the baseline level and slowly dropped back to the baseline level over the next few minutes during recovery.

**Figure 5:**
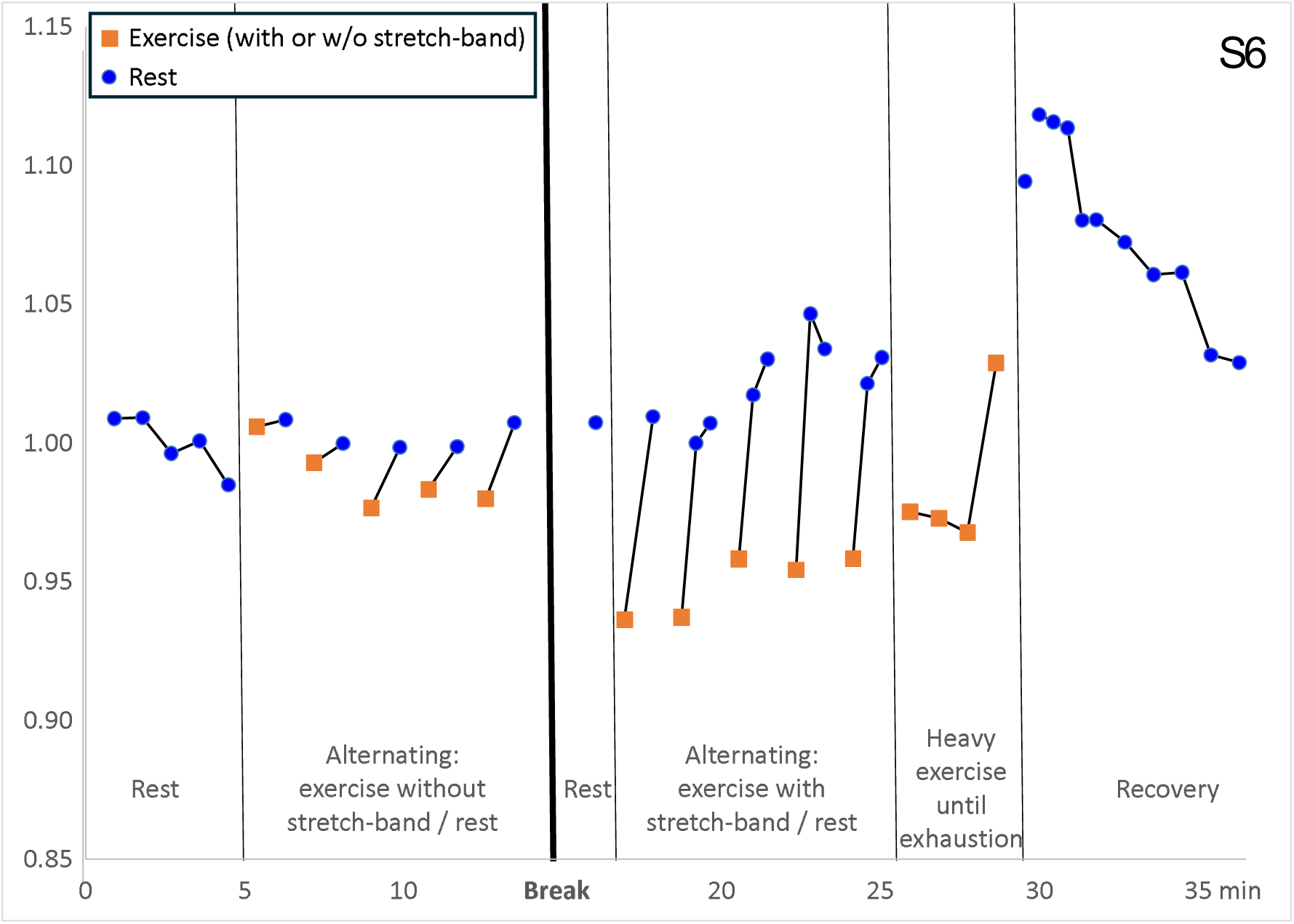
Time evolution of the fitted K peak-height for S6 during the different exercise phases with alternating exercise and recovery with and without stretch band and with exhaustive exercise according to the protocol shown in Fig. 2.

S7 performed isometric exercises, initially without the use of the stretch band, afterwards against the resistance. During the period without the stretch band (in which the leg was only lifted in the air), no significant change in the potassium signal in comparison with the baseline was detected (Fig. 6). In contrast, with the stretch band in place, the potassium signal dropped significantly by 3.1 % and returned to the baseline level at the termination of the exercise. However, at the high temporal resolution of only 6.75 s per spectrum, the SNR was low (which prompted us to overlay a moving average over the individual spectra in Fig. 6) possibly preventing the detection of a transition period.

**Figure 6:**
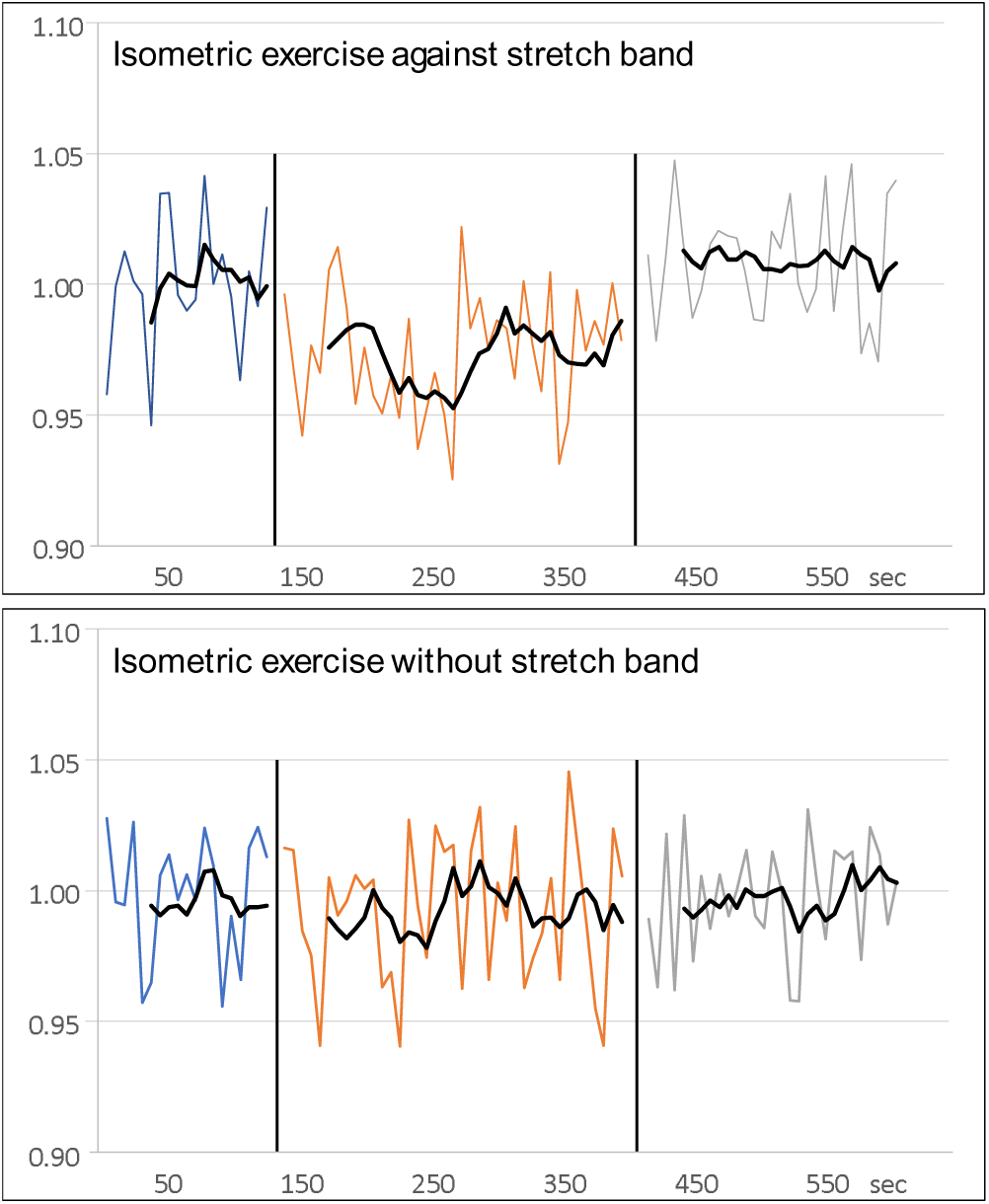
Time course of the fitted K peak-height for S7 during rest (blue lines), isometric exercise (orange), and recovery (grey) recorded with high temporal resolution of 6.75 s. Isometric exercise against resistance (top) and without resistance (bottom). Because of low SNR of the spectra, resulting in strong signal fluctuations, a moving average of 6 points is overlaid (corresponding to time resolution of 40 s).

For subject S8 MR-images of the resting and exercising thigh were acquired with high temporal resolution as described in the methods section to evaluate directly potential thigh movement and muscle contraction during lower leg exercise with and without stretch band. The sensitive area of the coil was estimated and for each of the acquired 100 MRIs per experiment with and without stretch band the enclosed muscle tissue area determined. Fig. 7a shows for both conditions a single MRI at rest and one example image during exercise showing greatest deviation from rest position.

**Figure 7:**
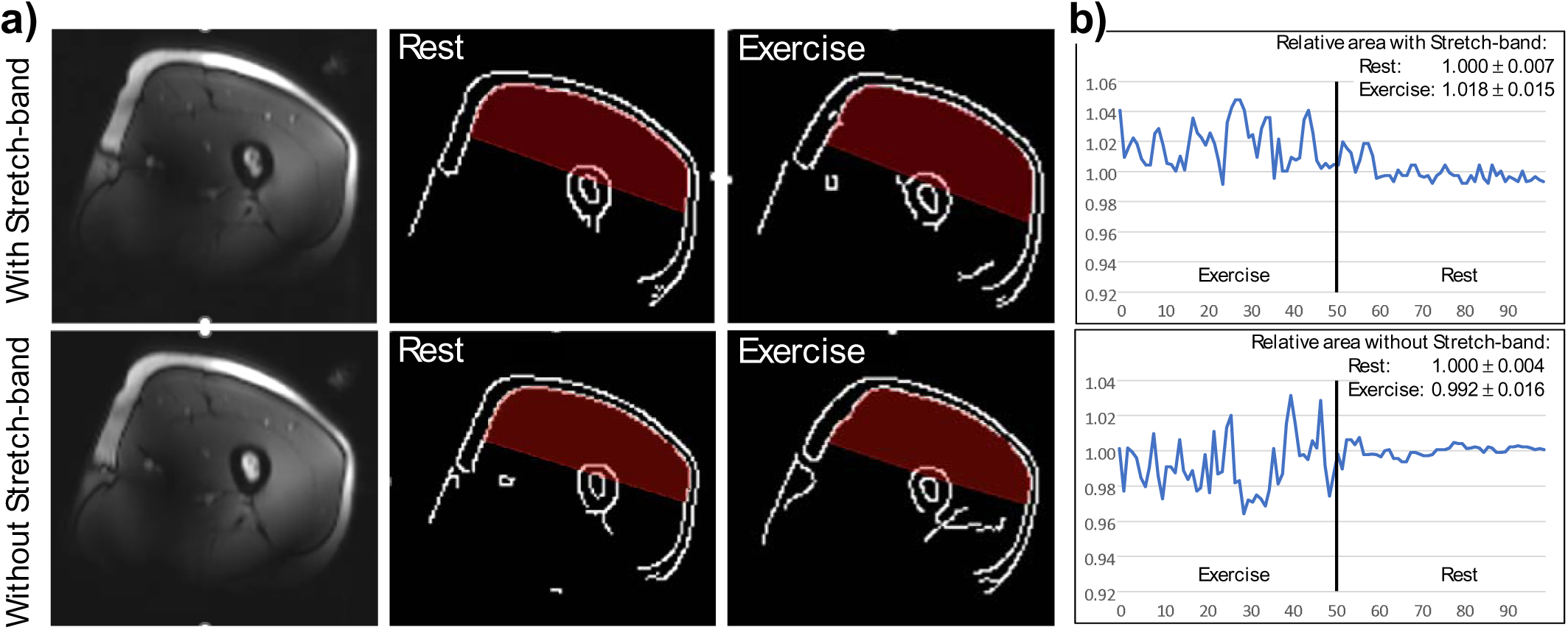
a) H-MR images at rest (left) with corresponding contour-plots (center) and example contour-plots during exercise (right) which showed greatest deviation from rest position. The tissue within the estimated sensitive region of the coil is overlaid in red. Top row: Exercise against resistance, Bottom row: Without movement without resistance. B) Analysis of the cross-sectional are within the sensitive volume of the coil for all 100 MRIs recorded during exercise and rest with (top) and without (bottom) stretch band.

Visually, the fluctuations appeared similar for exercise with and without stretch band (Fig. 7b). The quantitative results of this measurement showed that the area of the muscle cross-section in the sensitive coil volume fluctuated between -3.6% and +4.8% during exercise compared to rest. Nevertheless, the mean muscle area did not differ substantially: During the movement without the stretch band attached the muscle area was about 0.8% smaller than the mean during rest, while it was 1.8% larger during exercise with the stretch band compared to rest.

## Discussion

The main purpose of this study was to determine if changes in potassium concentration can be measured non-invasively in skeletal muscles using ^39^K-MRS during exercise, which is a precondition for potential clinical studies. It is well known that exhaustive exercise causes a decrease in intracellular potassium levels, as well as total muscle potassium content [14]. The aim was therefore to determine whether a decline in total muscle potassium content during dynamic knee extension can be discerned through the use of ^39^K-MRS. The main findings of this study were i) a consistent rapid decrease in total muscle potassium content in quadriceps muscles during moderate and heavy exercise of 5-6% returning to baseline during recovery; ii) these results are not explained by an altered muscle mass within the sensitive volume of the coil; iii) surprisingly, an initial potassium overshoot was observed during the recovery phase.

In the main experiment involving a standardized dynamic knee extension protocol with periods of rest, moderate and heavy exercise, and recovery we observed a significant decrease in total muscle potassium content in quadriceps muscles during moderate and heavy exercise in all participants consistent with results found in the literature. The average decrease of 5-6% was within the expected range based on previous invasive studies on biopsies [15,16]. In line with this finding, an increase in potassium content back to baseline levels was observed during recovery. This can be explained by the highly activated Na-K pump, which shifts the potassium from extracellular to intracellular and thus from the plasma to the skeletal muscle [18]. The adaptations of detected potassium levels to the onset and cessation of the exercise period appeared instantaneously without the detection of a transition period (on the timescale of our temporal resolution of about 1 min). While it is known that the potassium adaptation occurs rapidly especially upon commencement of exercise, this finding required further attention and prompted us to investigate possible artificial outcomes.

Although the quantitative potassium level changes during exercise were in the expected range, they may be due to technical issues, pretending intracellular potassium adaptations, i.e., leading to an artificial finding: Despite fixing the coil and the thigh, it may be possible that the coil changed its position in relation to the thigh during exercise, or that muscle contraction changed the cross-sectional area of the thigh, thus changing the muscle amount in the sensitive area of the coil. To investigate this issue, further exercise measurements were performed on three subjects.

The results of these additional measurements showed that dynamic knee extensions without resistance, i.e., performing similar lower leg movements as against resistance but with much lower power output, did not lead to significant potassium level changes. Also lifting the lower leg on a support so that the knee was at the maximal stretched position at rest did not lead to changes in detected ^39^K-levels. This indicates that the detected changes were not due to a changed coil position during exercise relative to the muscle. Furthermore, fast real-time MRI during rest and exercise revealed no significant change in the cross-sectional area of the tissue in the sensitive volume of the coil during exercise. The volume appeared slightly increased during movement against resistance compared to rest and compared to movement without resistance, thus indicating rather higher muscle tissue (and potassium) in the sensitive area of the coil during exercise.

Interestingly, a potassium “overshoot” clearly above baseline levels was detected in S6 (and slightly also in S1 and S3 in the main study) at the beginning of the recovery period after exhaustive exercise followed by a slow decrease to baseline. In this recovery period, the subject did not move and an artifact due to movement or cross-sectional area change can almost be excluded. A potassium overshoot was observed previously after exercise and was explained by ^39^K redistribution into the intracellular space after exercise exceeding baseline levels, but this was detected only 48 hours after exercise [13].

However, a potassium overshoot directly after exercise has not been described in the literature to the best of our knowledge and seems to oppose current physiological description of re-establishing the baseline potassium level after exercise (see introduction) [18]. Besides the origin of the determined potassium overshoot, the reason for the very rapid adaptation of the determined potassium levels after cessation of exercise also remains unclear, while a fast response of the potassium level upon onset of exercise has been described previously in invasive studies [15,16,18]. Overall, the measurements performed in addition to the main study suggest that the detected potassium level changes indeed reflect response to exercise and are not due to technically induced artifacts. However, further measurements are required, including measurements on more subject, to address the remaining uncertainties discussed.

Our study has some limitations: First, the exercise intensity was not quantified for this initial feasibility study. The individual and subjective exercise intensities may have contributed to the variance of potassium level adaptations. Therefore, the comparison with measurements without resistance proved very valuable. Quantification of power output during exercise is recommended for further studies. Second, localization of the spectra was only achieved over the sensitive area of the coil to allow high temporal resolution. Therefore, muscles that were less activated by exercise have likely contributed to the spectra, possibly reducing the detected adaptation of potassium levels. Enhanced localization may be used in further studies, but at the expense of longer measurement times. Third, no absolute potassium concentrations were determined, and only relative potassium changes are reported. Fourth, the study population is biased as most of the participants were male and participated regularly in sports, which has resulted in the development of a larger muscle mass than that of the average population. Nevertheless, this is unlikely to have had a significant impact on the results, as Nielsen et al. demonstrated that there is no difference in the release of potassium from working muscles between trained and untrained muscles [17].

## Conclusion

In summary, the findings of this study strongly suggest that intracellular potassium changes can be assessed in vivo using non-invasive MR methods at ultra-high field. The study also demonstrates that potassium levels in muscle can be monitored – though non-localized – in real-time during exercise. MRI techniques are non-invasive and do not rely on ionising radiation, which opens up the possibility of investigating potassium alterations in physiological studies or in disease.

## Data Availability

All data produced in the present study are available upon reasonable request to the authors

